# “Our kids are our future”: Barriers and facilitators to vaccine uptake and timeliness among Aboriginal children aged under five years in Boorloo (Perth), Western Australia

**DOI:** 10.1101/2025.01.08.25320235

**Authors:** Carla Puca, Paige Wood-Kenney, Naomi Nelson, Jordan Hansen, Judy Mathews, Erin van der Helder, Justin Kickett, Melanie Robinson, Katie Attwell, Anastasia Phillips, Valerie Swift, Christopher C. Blyth, Samantha J. Carlson

**Affiliations:** Wesfarmers Centre of Vaccines and Infectious Diseases, The Kids Research Institute Australia, Perth, Western Australia, Australia; Boorloo (Perth) Public Health Unit, North Metropolitan Health Service, Perth, Western Australia, Australia; South Coastal Babbingur Mia, Perth, Western Australia, Australia; Department of Infectious Diseases, Perth Children’s Hospital, Perth, Western Australia, Australia; Boorloo Community Member, Perth, Western Australia, Australia; Child and Adolescent Health Service, Perth, Western Australia, Australia; School of Social Sciences, The University of Western Australia, Perth, Western Australia, Australia; National Centre for Immunisation Research and Surveillance (NCIRS), Sydney Children’s Hospital, Sydney, New South Wales, Australia; School of Allied Health, Curtin University, Perth, Western Australia, Australia; School of Medicine, The University of Western Australia, Perth, Western Australia, Australia; Department of Microbiology, QEII Medical Centre, Perth, Western Australia, Australia

## Abstract

**Introduction:** Rates of several vaccine preventable diseases, and associated hospitalisation, are higher among Aboriginal and/or Torres Strait Islander children than non-Indigenous children. Western Australia has among the lowest childhood vaccine coverage in Australia, particularly among Aboriginal and/or Torres Strait Islander children. Delayed vaccination is also more common in this population. This project aimed to understand the barriers and facilitators to vaccine uptake and timeliness among Aboriginal and/or Torres Strait Islander children aged under five years in Boorloo (Perth).

**Methods:** The Tailoring Immunisation Programs method was used to determine the barriers and facilitators to vaccination. Between March – November 2023, in-depth interviews were conducted with 18 parents/carers of Aboriginal and/or Torres Strait Islander children in Boorloo who were currently or previously overdue for one or more childhood vaccines. Qualitative data were analysed in NVivo 14 using deductive and inductive coding following the Capability-Opportunity-Motivation-Behaviour model.

**Results:** The majority of participants believed vaccines are safe, effective and supported vaccination as a means to strengthen the next generation of Aboriginal and/or Torres Strait Islander children. Barriers to on-time vaccination related to access and ineffective reminder systems. Participants expressed limited knowledge about additional vaccines recommended for Aboriginal and/or Torres Strait Islander children under Australia’s National Immunisation Program.

**Conclusion:** An increase in vaccine coverage and timeliness will optimise protection and decrease the burden of disease in Aboriginal and/or Torres Strait Islander children in Boorloo. This can be supported by policy interventions to better cater for the needs of families, including addressing transport challenges, being mindful of the wider network of kin and care relationships, ensuring the funding of Aboriginal health services, and co-designing culturally appropriate resources. The federal government should use the Centrelink system to notify all eligible families of pending vaccines due.

## Introduction

The Aboriginal and Torres Strait peoples are the traditional owners of the land, sea, and waterways, and have lived in Australia for at least 65,000 years. There is great diversity between Aboriginal and Torres Strait Islander peoples in Australia, with over 250 Indigenous language groups, and around 800 dialects(1); each with their own culture, customs, beliefs and histories. Our research covers the area of Noongar Boodja (Country). The Noongar people are Aboriginal Australians from the south-west corner of Western Australia, having lived in this area for at least 45,000 years. There are 14 different Noongar clan groups within this area. Boorloo (the Noongar name for Perth) is the traditional lands of the Whadjuk Noongar People.

National data shows that rates of some vaccine preventable diseases (VPDs) including influenza, pertussis, invasive pneumococcal disease, meningococcal B and *Haemophilus influenzae* type b are higher among Aboriginal and/or Torres Strait Islander children than non-Indigenous children.(2) Childhood vaccination is an important and successful public health measure to reduce morbidity and mortality from such infectious diseases. Australia’s fully funded *National Immunisation Program* (NIP) offers free immunisation against 17 VPDs to all residents from birth to adulthood. Additional vaccines and doses are recommended and funded for Aboriginal and/or Torres Strait Islander children at certain ages.(3) These vaccine doses (hereafter, ‘additional vaccines’) are in addition to the routine vaccinations offered on the NIP, and include vaccines for meningococcal B and hepatitis A, plus additional doses of pneumococcal vaccine.(4)

*The National Immunisation Strategy 2019-2024* set targets of 95% immunisation coverage rates for children aged 1, 2 and 5 years old.(5) Coverage rates are measured using data from the Australian Immunisation Register, the world’s first electronic register, established in 1996.(6) Notification of an Aboriginal and/or Torres Strait Islander status on the register is voluntary. Aboriginal and/or Torres Strait Islander children aged 5 years were the first to achieve the 95% coverage target.(7) However, gaps in immunisation uptake and timeliness exist across the country. Immunisation coverage rates in Western Australia (WA) are amongst the lowest in the country for both Aboriginal and/or Torres Strait Islander children, and non-Indigenous children. In 2022, only 82.2% of Aboriginal and/or Torres Strait Islander children in WA were fully vaccinated at 2 years of age (national average 89.1%)(8), compared to 91.0% of all children in WA (national average 92.0%).(9) These rates have declined since the start of the COVID-19 pandemic.(10, 11)

Delayed vaccination is also more common among Aboriginal and/or Torres Strait Islander children than non-Indigenous children in Australia.(12, 13) The Australian Immunisation Register does not show why children receive vaccinations late, although reasons could include deliberate decisions by parents to wait until their children are older, or access and systemic barriers. Delay in receipt of one dose can drive further delays, with complexity added by the need to manage catch-up plans(12). Improving timeliness of vaccination is an essential element to improving coverage by 1 and 2 years of age.(12, 14), (15)

Families can access their children’s vaccinations at general practices, community immunisation clinics and Aboriginal Medical Services.(16) Routine and additional vaccines are free for Aboriginal and/or Tores Strait Islander children, but some private vaccination providers may charge a consultation fee for the visit. It has been well established that Aboriginal and/or Torres Strait Islander peoples experience access barriers to health care services, due to lack of cultural safety within mainstream health services, geographical isolation and financial barriers to accessing care.(17, 18) These may also impact timely access to immunisation services for children.

Several initiatives have been introduced by state and federal governments to increase childhood vaccination coverage. The national *No Jab No Pay* (NJNPay) legislation requires children in Australia to be up-to-date with routine vaccinations in order for parents to receive Child Care Benefit, Child Care Rebate and Family Tax Benefit payments from the Commonwealth Government.(19) Western Australia implemented complementary *No Jab No Play* (NJNPlay) legislation, which requires children in WA to be up-to-date with routine vaccinations to enrol in early childhood education and care services, pre-kindergarten and kindergarten services, unless exempted (Aboriginal and Torres Strait Islander children are exempt).(20) The additional vaccines recommended for Aboriginal and/or Torres Strait Islander children in WA are not required under the NJNPay and NJNPlay legislations.

Previous studies have identified barriers to influenza vaccine uptake(21) and motivations to seek COVID-19 vaccination(22) among Aboriginal and/or Torres Strait Islander adults; but no studies have specifically sought to identify, from the caregivers perspective, the barriers and facilitators to vaccine uptake among Aboriginal and/or Torres Strait Islander children in Boorloo, WA. To address this significant gap, we sought to understand the views of parents and carers regarding the barriers and facilitators to vaccination among Aboriginal and/or Torres Strait Islander children aged under 5 years in Boorloo. Building on the relationships we formed with members of the Community throughout the project, we anticipate developing tailored, culturally appropriate solutions based on our findings. We aim to increase vaccine uptake in this population and work towards closing the gap in Aboriginal and Torres Strait Islander health outcomes.

## Methods

### Study Design

The Tailoring Immunization Programs (TIP) method(23) developed by the World Health Organization, was used to determine the barriers and facilitators to vaccination, by identifying central themes related to vaccine access, attitudes, and knowledge. The theoretical model and framework used in TIP is based on the COM-B model(24), which is widely used for any behaviour across any environment and has been adapted for an immunisation behaviour specific focus. For any behaviour (B) to occur, such as vaccine uptake, an individual needs the capability (C), opportunity (O) and motivation (M) to do so.

Aboriginal Research Assistant (PWK) took a leading role in community consultation, project design and administration, data analysis and the provision of ongoing cultural oversight. Ethics approval was granted by the Western Australian Aboriginal Health Ethics Committee (HREC1181) and the Child and Adolescent Health Services Human Research Ethics Committee (RGS5459).

### Community consultation

Throughout 2022, we hosted three pre-study workshops with Aboriginal Elders, Parents/carers of Aboriginal and/or Torres Strait Islander children, and those working in Aboriginal health. These workshops sought to: 1) garner support for our project (and to learn if this was a topic that members of the Boorloo Aboriginal community wanted researched), 2) acquire feedback on our proposed methods, and 3) identify community members’ understanding of potential barriers to vaccination, thereby helping to develop the interview questions. A final workshop, conducted in April 2024, brought together representatives of the research team and the community, including research participants, to interpret and discuss the findings to assist in the development of this paper. (See analysis section.)

### Eligibility

Participants were eligible to participate if they were aged at least 18 years, and self-identified as being the parent or carer of an Aboriginal and/or Torres Strait Islander child aged under five years who was currently or previously overdue for one or more vaccines. ‘Overdue’ was defined as 30 days or more delay after the recommended age for each vaccine listed on the NIP Schedule (including the additional vaccines recommended for Aboriginal and/or Torres Strait Islander children in WA). Participants were included if they usually resided in Boorloo and could read and understand English. Parents/carers of Aboriginal and/or Torres Strait Islander children were not excluded if they did not identify as Aboriginal and/or Torres Strait Islander themselves.

### Recruitment

Participants were recruited from three health organisations: Perth Children’s Hospital (PCH), Boorloo (Perth) Public Health Unit (PHU) and South Coastal Babbingur Mia (SCBM). Recruitment methods were co-designed with each partnering organisation.

#### Perth Children’ Hospital

PCH is WA’s specialist paediatric hospital and trauma centre providing tertiary level care to people aged up to 16 years of age. Parents/carers who attended an outpatient clinic at the hospital for any reason were opportunistically recruited into the project by a health worker if they met the eligibility criteria.

#### South Coastal Babbingur Mia

SCBM is an Aboriginal health service that provides flexible and supportive services for the Aboriginal and/or Torres Strait Islander community in Boorloo’s southern suburbs. Services include health assessments, child health checks, pregnancy support, immunisations, and a weekly playgroup. Parents/carers attending the weekly playgroup were opportunistically recruited into the project by staff at SCBM if they met eligibility criteria.

#### Boorloo Public Health Unit

Boorloo PHU is a metropolitan-wide public health unit that offers immunisation support and a culturally-safe home-visit vaccination service for Aboriginal and/or Torres Strait Islander families. During routine telephone calls to book home visits, Boorloo PHU Aboriginal Health Liaison Officers (AHLOs) informed parents/carers about the study. If parents were interested in participating, AHLOs sought verbal consent for researchers to attend the at-home vaccination appointment. Researchers then provided more information about the project and obtained informed written consent prior to data collection occurring.

### Data collection

Data were collected via semi-structured, in-depth interviews. We sought to recruit 6-8 participants per partnering health organisation, with an overall aim of interviewing until thematic saturation was reached, which we anticipated would occur after 18-24 interviews. Three research team members were involved in data collection: PWK (Aboriginal), CP (non-Indigenous) and SC (non-Indigenous), who all identified as female. Interviews were conducted by a maximum of two facilitators and participants were not known to the facilitators. At times, people known to the participant were also present in the interview (e.g., their parent, partner or child/ren). Prior to starting the interview, facilitators described the background and purpose of the project, gained informed written consent and asked participants to complete a short demographic survey.

All interviews occurred face-to-face, ran for approximately 30 minutes and were audio-recorded. Interviews with participants recruited through PCH and SCBM occurred in a private clinic room at that service. For those recruited through Boorloo PHU, interviews occurred in the participant’s home or at community outreach events. After completing an interview, participants were provided with a $40 gift voucher to thank them for their valuable contribution to the project. Specific consent was sought by researchers to access AIR records of the child/ren in the care of interview participants to ascertain the number and type of vaccines that were overdue. This was an optional request, and those who declined were still able to participate and receive a gift voucher. After 18 interviews, thematic saturation was reached and recruitment ended.

### Interview guide

Following the COM-B model, we asked participants about their:

- Capability: knowledge, skills, and abilities to engage in vaccination.
- Opportunity: external factors that impact vaccination, such as access to services and vaccine information (physical opportunity) and the influence of family and friends on the decision to vaccinate (social opportunity)
- Motivation: attitudes, emotions, intentions, and beliefs that influence vaccine decision making and behaviour.

### Analysis

Demographic data provided by participants were analysed in Microsoft Excel using descriptive statistics. Socioeconomic conditions, based on postcodes provided, were assessed using SEIFA (Socio-Economic Index for Areas).(25) Postcodes were aligned to the decile score within the ‘Ranking Within State or Territory (Western Australia)’ outlined in the Postal Area (POA) Index of Relative Socio-economic Advantage and Disadvantage, 2016.

Qualitative data were analysed in NVivo 14 using deductive and inductive coding following the COM-B model. Data were individually coded by PWK and CP. A series of meetings were held between PWK, CP and SJC to discuss differences and similarities in coding and to finalise coding decisions. Quotes have been used throughout to illustrate themes. We elected not to assign pseudonyms to quotes out of concern that participants may be identifiable where more than one quote from a participant has been used.

Participants who consented to being contacted about research findings were invited to a Community Forum in April 2024, which was also attended by other Aboriginal and/or Torres Strait Islander community members. At this forum, the research team shared preliminary findings and sought verbal feedback on whether the findings resonated with attendees, and how the findings could be communicated about in most respectful way. Final themes presented in this paper incorporate the feedback received..

## Results

Interviews were conducted between March and November 2023. Of the 18 participants, six (33%) were recruited from PCH, four were recruited from SCBM (22%) and eight were recruited from Boorloo PHU (45%). Most were female (94%), aged between 25-34 years (56%), currently single (72%), not currently in paid work (72%), with an education level of grade 10 or equivalent (61%). More than half of them (61%) cared for two or more children under the age of five years. A full list of participants’ demographic characteristics is included in Table 1.

**Table 1:**
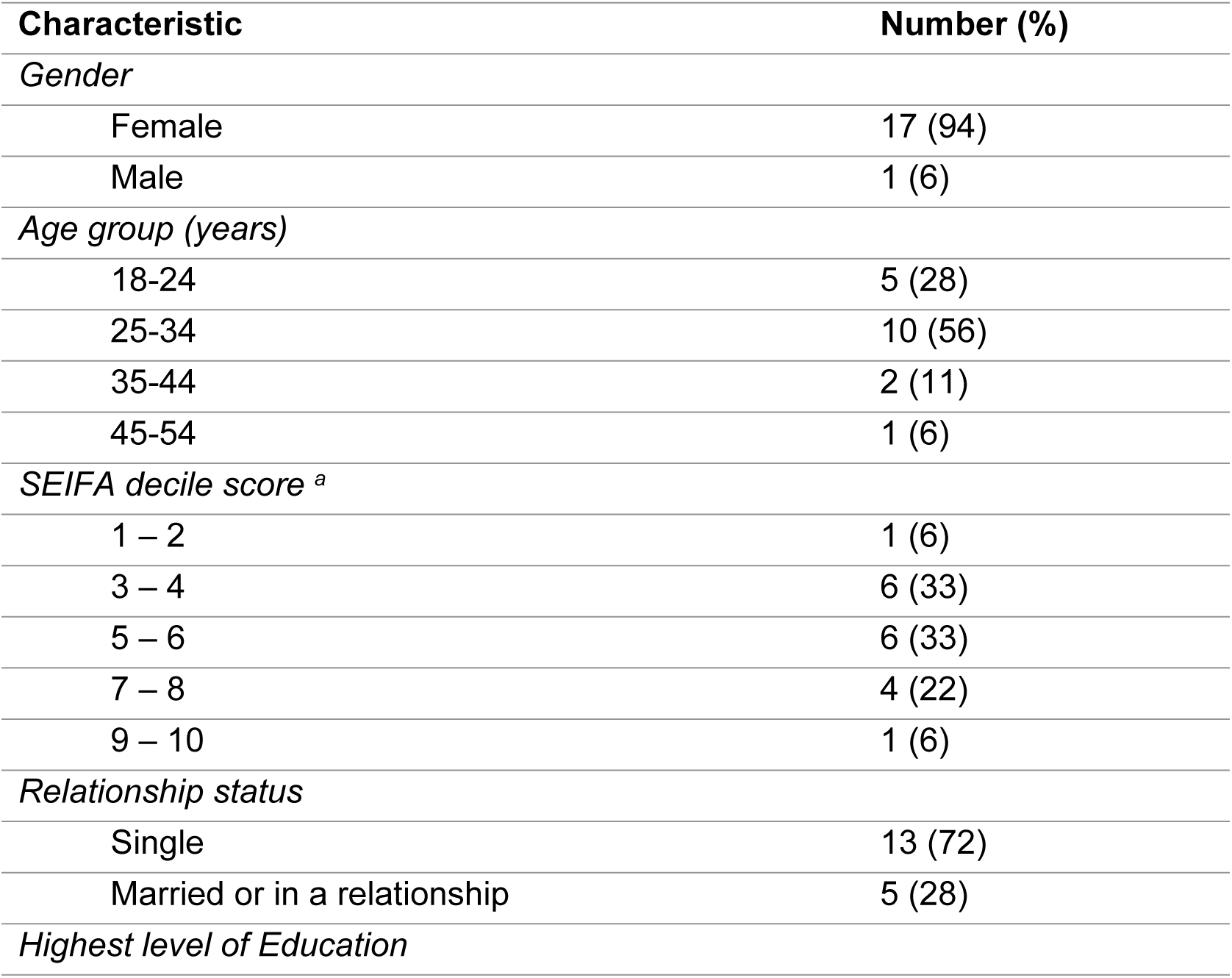
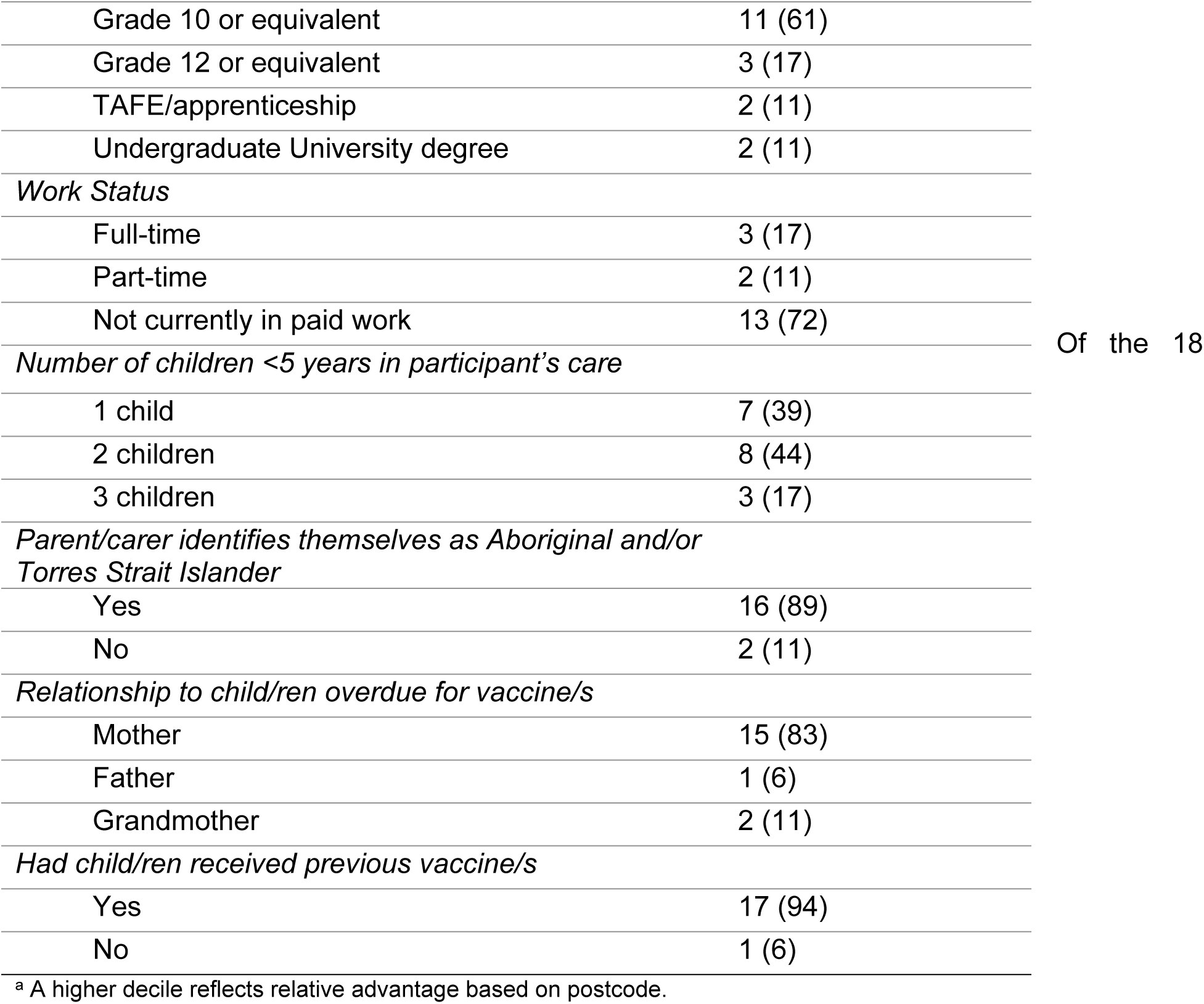
Demographic information of participants.

participants, 14 (78%) provided specific consent for the study team to access AIR records for any child they cared for under the age of 5 years (representing 22 children in total, aged between 4 and 55 months). Of the 22 children, 11 (50%) were up-to-date with NIP vaccines but overdue for the additional vaccines recommended for Aboriginal children, 10 (45%) were overdue for both NIP vaccines and additional vaccines, and 1 child was up-to-date with both (but is included in this study as their older sibling was overdue for one or more vaccines). Only one child (aged 7 months) had received no prior vaccinations.

### Facilitators to vaccination

#### Physical Opportunity - Benefit of culturally sensitive health and outreach services

Many participants spoke of the benefit of culturally sensitive health and outreach services in helping them access vaccinations for their child/ren. The provision of transport support (such as the free ‘pick up’ service offered by SCBM) assisted parents/carers in accessing vaccines for their child/ren.

> *I’ll ring [SCBM] and let them know that I think [my daughter] is due for her needles… and they’ll check and go, “Oh, yeah. She is. Do you want to arrange a time to come in? Do you need to be picked up, and we’ll pick you up and drop you off?.”*
>
> - Mum, 18-24 years

Participants also spoke favourably about at home immunisation services (such as the service provided by Boorloo PHU). These services helped them to access vaccinations in the comfort of their home.

> *I like how [Boorloo PHU] come out here…it’s a lot easier, and they just text when the kids are due [for their immunisations].*
>
> - Mum, 35-44 years

Some described their preference for using Aboriginal health services because of the welcoming environment. Some also described feeling more comfortable receiving immunisations from Noongar health workers.

> *It’s just easy and good here [at SCBM]. Here it’s good. It feels like home. You’re always welcomed. They have the playgroup here if I needed my other kids to get watched.*
>
> - Mum, 25-34 years

> *You can tell with the [Aboriginal Indigenous Health] team they know how to talk to Indigenous fellas, because with us it’s just talk to us like we’re people. You don’t have to talk to us like we’re slow or something…I think just because we’re Indigenous I think they – not all of them, just a few – if they haven’t dealt in the community before they think they have to deal with us in a certain way. I’m like, ‘Just talk to us like we’re human, we’re fine.’*
>
> - Mum, 25-34 years

#### Social Opportunity - Family and community influence

Only some participants spoke of a social expectation among their family and community to vaccinate their child/ren. This included being reminded by friends and family to vaccinate, and not being welcomed at community events unless *“you get [your kids] vaccinated”*

> *…The Elders will jump down your throat [about childhood vaccination]…[They say] “it just needs to be done.”*
>
> - Mum, 25-34 years

For most participants, however, vaccination did not appear to be an expectation within their family or community, nor was it commonly discussed in their social networks.

#### Motivation - Importance of vaccine information coming from trusted health authorities and community leaders

Participants had high trust in health care workers to deliver vaccine information and advice. However, there were mixed responses on whether they trusted the government’s advice about vaccinations. Some trusted this advice because childhood vaccines are *“standard for every kid”* (Mum, 25-34 years) while others felt strongly that the government “*lie about everything*” (Mum, 25-34 years) and *“will obviously say whatever they need to say to get people to do what they want”* (Mum, 25-34 years).

Those who mistrusted government indicated the importance of vaccine information and advice coming from trusted health workers and community leaders. Our only vaccine hesitant participant explained:

> *[Have vaccine information] come from a health care worker, come from Elders. Educate an Elder and say, “This is why you’re getting it done”. If they don’t believe it, then I’m not going to believe it.*
>
> - Mum, 25-34 years

#### Motivation - Strengthening the next generation

Some participants described wanting to “*protect*” (as described by Mum, 25-34 years) their children from illness and strengthen the next generation of Aboriginal and/or Torres Strait Islander children as a primary driver to vaccinate. As put simply by one participant: *“Our kids are our future”* (Mum, 25-34 years).

> *I’ve got a very strong…community-based family at home, especially when it comes to me [and] my baby…I guess they just want…all our Aboriginal babies to be healthy…so immunisations especially in our family are a must…We just want our generation or the next generation to be stronger and better than we are at the moment…The [babies are] very strong and they’ve got a lot of value for immunisations…*
>
> - Mum, 25-34 years

This motivation to vaccinate was mirrored even among participants who were unsure about the effectiveness of vaccines. Furthermore, recommendations from trusted health workers helped to bridge the gap where doubts lay.

> *I can’t really say [whether vaccines work]… I don’t know if [vaccines] help or not. But…if I’m being told [by a health worker that] it does help, then obviously I’m going to do the right thing and protect my kids from certain [illnesses] that they could possibly get.*
>
> - Mum, 25-34 years

#### Motivation - Routine vaccines viewed as safe

All but our vaccine-hesitant participant perceived routine vaccines as safe. They put their trust into routine childhood vaccines because of how long they have been in use, and felt there was sufficient and acceptable data on any possible short-term and long-term side effects.

> *The immunisations have been around for years since I was a kid, since my mum was a kid, and we’re all okay.*
>
> - Mum, 25-34 years

Participants also perceived routine childhood vaccines as safe through their lived experience: their child/ren had not experienced any major adverse reactions to vaccines. However, some raised that their view on vaccine safety would likely change if their child/ren ever experienced a major adverse reaction.

One participant had concerns about why live-attenuated vaccines were needed. However, our AIR check indicated that this participant’s child had previously received live attenuated vaccines on the NIP. This indicates that her concerns were not strong enough to motivate her to refuse them.

> *I guess [vaccines are safe]…it’s just I don’t know [about] live vaccines…Why are [my children] getting injected with a live vaccine to prevent them from getting this disease? Why don’t you just make something to stop them from getting the disease instead? This whole thing confuses me.*
>
> - Mum 25-34 years

#### Motivation - Routine vaccines viewed as effective

Most participants believed that vaccines are effective in preventing illness and associated hospitalisation in children.

> *I do believe [vaccines work]….it gets rid of all the viruses, … if [children] didn’t have their needles….Perth Children’s Hospital would have been really decked out with sick kids.*
>
> - Grandmother, 45-54 years

A minority of participants were unsure of the effectiveness of vaccines as their child/ren has been sick despite being previously vaccinated. These participants held the misconception that routine vaccinations prevent all illnesses in children.

> *I don’t know if [vaccines] help or not…you’d think [kids]… wouldn’t get as sick as often [if they’re vaccinated]… but my oldest daughter, she’s been immunised when needed, but she’s the one who’s always been sick, always in hospitals. So, I’m not sure if [vaccines work] – I know they probably do work, but I don’t know.*
>
> - Mum, 18-24 years

#### Motivation - Intention to catch-up

At the time of interview, all participants described their child/ren as being caught up with their vaccines or described an intention for their child/ren to be caught up. Those intending to catch up had plans in place for the child’s upcoming vaccinations.

> *Yeah. I’ve got him booked in [to be vaccinated] next Friday*
>
> - Mum, 25-34 years

#### Capability-Knowledge about vaccines and how to access them

Participants frequently mentioned meningococcal vaccines. Many recalled media campaigns observed in their childhood featuring images of the long-term impacts of a meningococcal infection and promoting the importance of vaccination. Participants did not express detailed knowledge about any other specific vaccines, or the diseases they protect against. Despite this, there was an overarching understanding among most participants that the diseases these vaccines protect against could be life-threatening to their child/ren.

> *They [diseases that the needles protect bubs against] could be serious and kill them. That’s why we have the vaccinations, to try and prevent that happening.*
>
> - Mum, 25-34 years

Participants were also aware that the parent-held child health record book provided by the Western Australian government health system contained a list of vaccines their child needed, and the ages at which each vaccine was recommended. However, some had since lost their record book, or it had been accidentally destroyed.

Participants were informed of possible vaccine side effects at the time of vaccination by health care workers. They reflected that they could minimise these side effects by administering paracetamol before vaccination or in the hours following vaccination. Awareness of possible side effects did not appear to deter parents from choosing to vaccinate their child/ren. However, one participant held the misconception that convulsions can be caused after any vaccine if paracetamol is not administered prior to vaccination.

Finally, there was high awareness among participants of where they could go to get their child/ren vaccinated. These included their local GP clinic and Aboriginal health and medical services in Boorloo.

### Barriers to on-time vaccination

#### Physical Opportunity – difficulty accessing health clinics

##### Lack of transport options

Many participants described access to health services as being a key barrier to on-time vaccination. Access barriers were primarily associated with difficulties in travelling to health services to receive vaccinations, due to a lack of transport options. Some of these participants did not have their driver’s license and/or access to a car, and therefore were reliant on public transport; this was particularly challenging for parents/carers with multiple young children.

> *I don’t have a car or anything…no license, and I’ve only got a double pram for the three kids.*
>
> - Mum, 25-34 years

These participants described how having access to a car would be make it easier to get their child/ren’s vaccinated on time.

##### Managing child and family responsibilities

Several of the participants shared stories about the significant, conflicting personal and family challenges they had experienced, making it more difficult to manage child and family responsibilities, including obtaining vaccinations. Some of these conflicting challenges included death of a close family member, overcoming personal physical and/or mental health issues, supporting a family member who was experiencing problems related to the use of alcohol and/or illicit drugs, frequently moving between multiple residences, and supporting a family member who was experiencing intimate partner violence.

One participant described the unexpected passing of her husband a few months prior to the interview. This had a devastating impact on her mental health and her ability to undertake many of the parenting tasks she did before her husband passed:

> *I’m usually pretty good with it all. But this is where it gets tough. Her [daughter] father [and my husband] passed away [3 months ago]. So I’ve been staying home….I’ve been finding it hard…When I get into the car, I have panic and anxiety attacks, which I’ve never ever experienced in my life. So I’m only behind with her [needles] because I’m still learning to [cope].*
>
> - Mum, 25-34 years

Others spoke of the challenges they had in caring for multiple children, sometimes without support from a partner, while balancing work and other family responsibilities. These pressures made it difficult to vaccinate their child/ren on time.

> *With my first child, I used to keep track of [vaccinations] all the time myself. As the second one came, I would try to keep track of it, but sometimes I forget. With this one, the third one, I’ve just not been able to be on top of it.*
>
> - Mum, 18-24 years

#### Physical Opportunity – ineffective reminder systems

Participants were predominately made of aware of vaccine due dates through reminder systems that form part of Australia’s vaccination governance system. However, most participants only received such reminders once their child was already overdue. Reminders predominantly came from Centrelink (a Commonwealth Government agency that provides income support to eligible Australians) informing parents/carers that their child did not meet their immunisation requirements, as per the *No Jab, No Pay* legislation. The notifications informed them their Family Tax Benefit payment would be reduced (and their Child Care Subsidy stopped, for those who send their child/ren to early childhood services). Participants considered these notifications useful as a reminder to vaccinate their child, but they also regarded them as threatening.

> *It’s kind of like a threat, almost, the way that [Centrelink] notify you [that your child is overdue for a vaccine], because they say, ‘If you don’t get it done we’ll cut off your [Family Tax Benefit] payments’, so they put the pressure on you a little bit.*
>
> - Dad, 25-34 years

A few participants spoke about receiving reminders from other sources, such as phone calls, emails and/or notifications in the mail before routine vaccines were due. However, these participants had an existing relationship with a health service or local clinic, which was the source of the message. Some of these participants also noted that even though they were linked in with a health service, they were not notified at any time about the additional vaccines recommended for Aboriginal and/or Torres Strait Islander children. Meanwhile, participants who were not linked in with a health service did not appear to receive vaccine reminders prior to due dates.

#### Physical opportunity – limited access to vaccine information presented in user-friendly format

Most participants described not receiving previous information or resources about childhood vaccination. A small number had received information pamphlets about vaccinations in hospital shortly after their child was born, but some did not find these particularly useful at that juncture.

> *I just don’t have the time [to read immunisation pamphlets] juggling with a new baby and all that.*
>
> - Mum, 18-24 years

Participants who had an existing relationship with a health service also described receiving information pamphlets about vaccinations during visits. However, some described these resources as difficult to understand, and believed that face-to-face conversations would be more beneficial. They also highlighted the need for vaccine information to be presented in simpler terms, without the use of medical jargon.

> *Some people don’t know how to read and write properly. If they had the person sit down with them and talk to them about their kids and what [vaccines] they’re getting done and that, that would be more helpful for the Aboriginal community.*
>
> - Mum, 18-24 years

#### Capability – Knowledge

##### Unaware that child is due for vaccines

Above, we described our participants’ experiences with inadequate reminder systems that did not occur at all, did not call their child in for the additional vaccines, or (in the case of Centrelink) came too late in the form of notifications threatening the cancellation of financial support. Without such reminders, there was little to prompt participants to get their children vaccinated on time, because most described not knowing when their child was due for vaccines.

> *To me, I didn’t really know much about vaccine dates. I obviously knew they had to happen but I didn’t know when exactly they had to happen, what age, what months.*
>
> - Dad, 25-34 years

However, most participants had high awareness of where they could go to receive vaccines for their children.

##### Limited knowledge about additional recommended vaccines

Only a few of our participants were aware of the additional vaccines recommended for Aboriginal and/or Torres Strait Islander children. They tended to hold misconceptions on the reasoning behind the recommendation, which they often associated with physiological differences between Indigenous and non-Indigenous bodies. Some believed Aboriginal and/or Torres Strait Islander people had a *“weakened immune system”* (Mum, 18-24 years) or that there were *“genetic reasons behind it”* (Mum, 25-34 years) or that *“….[Aboriginal] DNA…and blood is constructed differently”* (Mum, 25-34 years).

Some also raised fears that appeared to reflect Australia’s history of colonisation and genocide of its Aboriginal and/or Torres Strait Islander peoples.

> *I don’t know. Why do they got to get them? That’s what I’m always confused. … is something wrong with us that we’ve got to get additional protected?… Why have we got to get that and no-one else does? Or are they trying to kill us off or I’m like what’s going on here?*
>
> - Mum, 25-34 years

Most of our participants learned about the availability of additional vaccinations through the interviews. We took this opportunity to explain the rationale behind them, and how this connected to risk factors in the community rather than differences between bodies. Following these conversations, which arose organically during the interviews, almost all participants indicated their desire for their child/ren to receive the additional vaccines.

#### Capability – Child or parent unwell at time of scheduled vaccine

A small number of participants described themselves or their child/ren being unwell at the time of scheduled vaccines, as the reason for being overdue.

> *I was a bit late for the last two’s immunisations, yeah, because my boy was sick. He had like a sinus infection, his ears were a bit runny, poor thing. So he was a bit late, a few months behind for his needles. My daughter, she had a fever the day I took her in [to get vaccinated]….and they said” No, we can’t [vaccinate her] while she’s sick.’…if they’ve got a little bit of a fever [the GP won’t vaccinate them].*
>
> - Mum, 25-34 years

### Suggested Strategies

We asked participants how governments and health services can help parents like themselves vaccinate their children on time. Here, we provide their suggestions:

#### Improved reminder system

Participants were in agreeance that receiving a reminder notification one to two weeks before vaccines are due – rather than only receiving a notification once already overdue – would make it easier to vaccinate their child/ren on time. Some suggested these reminders could come in the form of an email, text message or notification from MyGov (the secure, online platform used to access Australian government services, including Centrelink).

> *It will be helpful if they…send a reminder to my phone, like a text message or a thing on my MyGov where I normally go put my [Centrelink] form in and so I’ll end up checking it. And that’ll actually work a lot better, where I can plan a day to go out to get the needles done.*
>
> - Mum, 18-24 years

#### Improved access to vaccination services

Improved access to vaccination services would also assist parents/carer in getting their child/ren vaccinated on-time. Participants suggested a ‘pick up’ service, where transport to and from a vaccination service can be provided for parents/carers and their child/ren, and a ‘home visit’ service, where vaccinations can be provided in the home. Some also suggested ‘pop-up’ vaccination clinics at community centres and shopping centres, inspired by the services that they had observed during the pandemic:

> *…when the COVID vaccine got rolled out, they had all the pop-up clinics everywhere and then you just went and sat. But there were quite a few people, so you got through quite quickly. You could probably do like similar, just on child vaccinations.…a drop in [service]…at a community center or shopping centers… It could even be, like, in a clinical van that is outside and it can just be either a waiting…You could put your number down, go do your shopping and when they call you, be, like, ‘Hey, you’re up in line,” instead of having to wait there.*
>
> - Mum, 25-34 years

#### More appropriate information

Participants sought more information about childhood vaccinations to help inform vaccine decision-making. They were particularly eager for more information on the additional vaccines recommended for Aboriginal and/or Torres Strait Islander children. They wanted to know *which* additional vaccines are recommended, *when* they are due, and *why* they are recommended for Aboriginal and/or Torres Strait Islander children.

Participants also wanted this messaging to be easier to understand. They suggested that it focus on which diseases each vaccine protects against, the severity of these diseases, and why these vaccines are important.

> *[I would like] more of a breakdown of …not of what actually goes in the immunisations, but how it works around [my child] as he grows, if that makes sense…I don’t know much about needles in general or anything, but I know he has to have his hep B…and there’s other types of needles he’s got…I know that he’s got to get his needles, but it’s like, maybe breaking it down more on why. Like, what’s it useful for?*
>
> - Mum, 25-34 years

In terms of the medium for this messaging, most participants suggested pamphlets or brochures. Some suggested audio-visual resources could be helpful to deliver vaccine information to parents/carers. Importantly, participants emphasised trusted health workers and community leaders were the right people to deliver this information. They also suggested that conversations with healthcare workers should accompany and buttress any information resources provided.

In terms of timing, participants suggested that the best times to receive information on childhood vaccines were both during and after pregnancy.

## Discussion

To the best of our knowledge, this is the first study to identify the facilitators and barriers to vaccination of Aboriginal and/or Torres Strait Islander children in Boorloo, WA, from the parent/carer perspective. Our findings contribute to a body of research that recognises the need to understand why people *accept* vaccines, as well as why they delay or refuse them.(26, 27) Our participants were eager to vaccinate as a means to strengthen the next generation of Aboriginal and/or Torres Strait Islander children and all intended to ensure that their children eventually received all recommended vaccines, including the additional ones that many only learned about in conversation with us. Despite their willingness, however, participants faced key physical opportunity barriers. Access barriers stemmed from managing child and family responsibilities, and lack of transport options. This is in line with other TIP studies conducted in Australia, which also identified access barriers as some of the primary barriers to on-time vaccination(28–30). Prior research conducted by members of this team with government health service providers in country Western Australia similarly uncovered access barriers and transport challenges amongst the Indigenous and non-Indigenous families they serve.(31)

Lack of transport options might seem to be a simple concept to understand as an access barrier, and can certainly be addressed with appropriate resourcing. However, there are greater complexities in understanding how conflicting challenges impact how parents and carers manage child and family responsibilities, and how this can disrupt their access to vaccine services. Aboriginal and Torres Strait Islander people involve extended family and community members in childrearing, fostering a sense of shared responsibility.(32) This differs to the nuclear family-focused approach of non-Aboriginal parenting, where parents are the primary caregivers of children. The conflicting personal and family challenges experienced by some of our participants hindered their ability to vaccinate their child/ren on time. Historical and ongoing impacts of colonisation, racism and discrimination continue to impact health and social disparities among Aboriginal and Torres Strait Islander peoples(33, 34), contributing to these conflicting challenges. The on-going strength, resilience and political will of Aboriginal and/or Torres Strait Islander peoples in sustaining 65,000 years, the historical and ongoing effects of colonisation and racism have contributed to current inequities in the health and wellbeing of Aboriginal and/or Torres Strait Islander peoples. Aboriginal and/or Torres Strait Islander peoples have higher exposure to trauma such as physical and/or sexual violence, and unexpected death of a loved one, compared to the non-Indigenous population.(35) Aboriginal and/or Torres Strait Islander peoples also more frequently engage in harmful health risk behaviours, such as misuse of alcohol and drugs(36), experience higher prevalence of suicide(37), and have poorer overall health and social outcomes compared to non-Indigenous people.(38) The cumulative effect of these significant challenges create complexities in managing child and family responsibilities, subsequently making it difficult to access on-time vaccination. Governments and services must consider these conflicting challenges – experienced not only by parents, but also other family and community members involved in childrearing – as they collaborate with the communities they serve to design activities that support timely vaccine access.

Culturally safe and flexible vaccination services can help to overcome access-related barriers.(28) Participants suggested that some of these access barriers could be addressed by providing transport support to get to vaccination services, and improving access to culturally safe ‘home visit’ services. Participants also spoke of their preference for receiving immunisations from Aboriginal Health Workers (AHWs), Aboriginal Health Practitioners (AHPs), and general practitioners who identify as Aboriginal and/or Torres Strait Islander. We support the role and contribution of specific Aboriginal-led services, but deficiencies in mainstream programs may also be shaping this preference. A survey of staff working in Boorloo general practices in 2018 found that two-thirds of employees reported having received no cultural safety training. Staff also felt that the leadership within general practices did not prioritise ensuring high vaccination rates among Aboriginal and/or Torres Strait Islander children in their practice.(39)

We hope that continuing demand for Aboriginal-led services can be met following recent policy and practice developments. Previously, families’ access to AHWs and AHPs in Boorloo who could administer vaccines was limited, and vaccine administration could only occur under medical supervision.(40) However, as of April 2024, AHPs in WA who have undertaken immunisation training, and who are working in public clinics/services or a health service that is a member of the Aboriginal Health Council of WA, can administer immunisations independently.(41) This is a significant milestone towards improving culturally safe access to vaccinations for Aboriginal and/or Torres Strait Islander people in WA. Governments should ensure continued funding and support for these programs and services to improve access to timely vaccination for Aboriginal and/or Torres Strait Islander children in Boorloo.

Another major physical opportunity barrier identified by our participants was the (non) use of vaccine reminder systems. Participants deemed Centrelink prompts as ineffective, as they predominantly notified families about vaccinations once their child was already overdue. Non-indigenous participants in a study conducted almost a decade earlier used near identical language to describe the “rude” letters “saying you are going to be cut off.”(42) Australia’s immunisation register has the capacity to identify when a child is *due*, not just *overdue*. This is a missed opportunity to use existing infrastructure to help the parents and carers of *all* Australian children stay up to date with their vaccinations. Reminders are also important from a social justice perspective given that families face financial penalties for failing to vaccinate their children on time.

The federal government can improve its existing reminder and recall system to provide notifications with ample time before vaccines are due can help parents/carers to plan for vaccination appointments in advance. This is likely to be effective for the cohort we studied, since evidence demonstrates that notifying families of Aboriginal and/or Torres Strait Islander children prior to vaccine due dates via SMS reminders(43–45) or a telephone call(14), improves timely vaccination. Other health services are already leading the way. Boorloo PHU initiated a pre-emptive reminder and support program in April 2023(44), based on a New South Wales (NSW) program(14), to enable families of Aboriginal and/or Torres Strait Islander infants to commence timely vaccination. Importantly, the program provides practical support (including appointment bookings and home visits (as needed), in addition to reminders, from 5 weeks of age. Indeed, it is through this effective service that we recruited and interviewed some of our participants whilst their children received or caught up with their vaccines.

Being unaware of the additional vaccines recommended for Aboriginal and/or Torres Strait Islander children in WA was a key capability barrier among participants. Children can be classified as ‘up to date’ on AIR even if they have not received these vaccines, since the additional vaccines are not used in the assessment of overdue status. Several levels of intervention can be helpful here. First, health providers need further education and/or prompts to ensure that they ask all parents/carers whether their child identifies as Aboriginal and/or Torres Strait Islander. Second, this education should help HCWs to understand that additional vaccines are not included in a child’s ‘up to date’ status on AIR, and that a discussion with parents about these recommended vaccines should occur even for Aboriginal and/or Torres Strait Islander children who are otherwise fully vaccinated. Third, strategies are needed to help health providers discuss the rationale behind the additional recommendations for Aboriginal and/or Torres Strait Islander children with parents/carers. Since the rationale for these vaccinations is important to families, information about additional vaccines should identify the diseases that these vaccines protect against, and make the case why they are important for the next generation of strong Aboriginal children. It may also be worthwhile addressing misconceptions about differences between bodies of Aboriginal and non-Aboriginal children. The rationale for the additional vaccines should be conveyed carefully to avoid blaming or stigmatising Aboriginal and/or Torres Strait Islander people and to build on the strengths-framing message that we identified in this study: vaccines help to grow strong Aboriginal children. Vaccine information and educational resources that help to advance this message should be co-designed with community so that information is relatable and easy to understand. Our participants also emphasised that, where possible, these resources should be accompanied by a conversation with a health care worker about the benefits of vaccination.

We did not identify significant vaccine hesitancy among our participants; however, this may be due to how we sampled. Participants recruited through Boorloo PHU home-visiting service were clients of that service who are known to not be hesitant. Moreover, although we heard from a range of families living across the metropolitan aera, our findings do not represent all the voices and perspectives of all Aboriginal and/or Torres Strait Islander families in Boorloo. To mitigate these limitations and to validate the barriers and facilitators of vaccination we identified, we plan a second stage of quantitative work. We will use the findings in this study to develop and distribute a survey to a representative sample of parents/carers of Aboriginal and/or Torres Strait Islander children in Boorloo investigating barriers and facilitators for vaccination amongst a wider cohort of families.

Another limitation of the study was that it was not possible for an Aboriginal research team member to attend every interview, due to circumstances beyond the control of the research team. As such, participants may not have shared their views and experiences as openly with the non-Indigenous research team members. However, given the deeply personal stories that participants did share with the research team, we believe that we were the beneficiaries of a sense of trust from the beginning of each interview. This is likely due to the already established and strong relationships between the organisations we recruited through and their clients.

A major strength of this study was the contribution made by Aboriginal investigators as well as members of the Aboriginal community in Boorloo – their wisdom informed the study design and conduct, and the interpretation of data. The collaborative interpretation of findings between the research team and a community forum was a highly fruitful and rewarding activity, and is considered best practice for the conduct of Aboriginal health research by The Kids Research Institute Australia.(46) We hope that other research teams can benefit from this form of engagement.

## Conclusion

Parents and carers of children under five who had previously been late for or missed vaccinations were nevertheless supportive of vaccination. They saw immunisation as a means to strengthen the next generation of Aboriginal and/or Torres Strait Islander children and spoke of their preference for receiving immunisations from Aboriginal health workers and services. The major barriers to on-time vaccination related to access. Managing hardships experienced by immediate and extended family members involved in childrearing, as well as lack of transport options, impacted on timely access to vaccination services. Ineffective or non-existent reminder systems and limited knowledge about additional vaccines recommended for Aboriginal and/or Torres Strait Islander children were also identified as barriers to on-time vaccination. Program actors should consider these barriers and facilitators in co-designing programs and resources to increase vaccine uptake and timeliness in this population. Policy interventions to better cater for the needs of families can address transport challenges, consider the wider network of kin and care relationships, ensure the funding and growth of Aboriginal health services, and facilitate the co-design of culturally appropriate resources. The federal government should use the Centrelink system to notify all eligible Australian families of pending vaccines due.

## Data Availability

The data generated and/or analysed during the current study are not publicly available. Approval has not been given by the Child and Adolescent Health Service Ethics Committee and WA Aboriginal Health Ethics Committee to share data, even in partially redacted form. Our participants were informed prior to participation that their data would be kept in strict confidence. Given the confidential nature in the data they shared, the relatively small number of individuals that participated in the in-depth interviews, the potential for reidentification and the impact that this would have in community confidence in both research and immunisation, we are unable to provide deidentified or redacted data for publication.

## Author contributions

- **Conceptualization:** Naomi Nelson, Melanie Robinson, Anastasia Phillips, Valerie Swift, Christopher C. Blyth, Samantha J. Carlson
- **Data curation:** Carla Puca, Samantha J Carlson
- **Formal analysis:** Carla Puca, Paige Wood-Kenney, Naomi Nelson, Erin van der Helder, Justin Kickett, Katie Attwell, Anastasia Phillips, Valerie Swift, Christopher C. Blyth, Samantha J. Carlson.
- **Funding acquisition:** Naomi Nelson, Melanie Robinson, Anastasia Phillips, Valerie Swift, Christopher C. Blyth, Samantha J. Carlson
- **Investigation:** Carla Puca, Paige Wood-Kenney, Samantha J. Carlson,
- **Methodology:** Carla Puca, Paige Wood-Kenney, Naomi Nelson, Jordan Hansen, Judy Mathews, Anastasia Phillips, Christopher C. Blyth, Samantha J. Carlson
- **Project administration:** Carla Puca, Samantha J. Carlson
- **Resources:** Christopher C. Blyth, Samantha J. Carlson
- **Supervision:** Valerie Swift, Christopher C. Blyth, Samantha J. Carlson
- **Validation:** Carla Puca, Samantha J. Carlson
- **Visualization:** Carla Puca
- **Writing – original draft:** Carla Puca
- **Writing – review & editing:** Carla Puca, Paige Wood-Kenney, Naomi Nelson, Jordan Hansen, Judy Mathews, Erin van der Helder, Justin Kickett, Melanie Robinson, Katie Attwell, Anastasia Phillips, Valerie Swift, Christopher C. Blyth, Samantha J. Carlson.

## Acknowledgements

We extend our deepest thanks to our participants for openly sharing their stories, experiences, time, and for some, their homes, with us. We thank all who attended the Community Consultation workshops for sharing their knowledge and input. We thank Brydie Campbell, Bridie Watson, Anita Campbell and Maria Xaver for assisting with PCH participant recruitment; Emily Garlett for assisting with SCBM participant recruitment; and Danielle Nelson and Rebecca Labruyere for assisting with Boorloo PHU participant recruitment. We thank Louisa Paparo and Cathy Pienaar for assistance with data acquisition. We would like to acknowledge the contributions made by Fred Penny and others in attendance at the Community Feedback Forum who assisted in interpretation of findings. We thank the Kulunga Aboriginal Research Unit for assistance with ensuring our research responds to community needs and meets the standards for Aboriginal Health Research. This work was funded by the Communicable Disease Control Directorate, WA Health. CCB is supported by a NHMRC Investigator Award. SJC is supported by a Brightspark Fellowship.

## Notes

### Competing Interest Statement

The authors have declared no competing interest.

### Funding Statement

Yes

### Author Declarations

Ethics approval to conduct this study was granted by the Western Australian Aboriginal Health Ethics Committee (HREC1181) and the Child and Adolescent Health Services Human Research Ethics Committee (RGS5459). Informed consent was obtained from all participants prior to participating in the study. We informed each participant of the purpose of the study, their rights, and how to withdraw the study should they wish to. Participants were assured that their anonymity and confidentiality would be strictly protected, and that their data would not be made available to people outside of the research team. The consent covered participation in the study, the use of collected data for research purposes, and permission to publish the findings. This research complies with all relevant ethical guidelines, including the institutional protocols for research involving human participants and the Declaration of Helsinki.

